# PPRS-ID: Indonesian-Adjusted Partitioned PRS for Type 2 Diabetes using Obesity PRS Integration and West Javanese Population LD Mapping

**DOI:** 10.64898/2025.11.28.25341222

**Authors:** Kezia Irene, Jocelyn Verna Siswanto, Belinda Mutiara, Eugene Vito Sebastian, Jonathan Susanto, Restu Unggul Kresnadi

**Affiliations:** PT Kalbe Farma Tbk, 10510 Central Jakarta, Indonesia; Kalgen Innolab Clinical Laboratory, Jakarta, Indonesia

**Keywords:** Cross-Population Generalizability, Polygenic Risk Score (PRS), Genome-Wide Association Study (GWAS), Ancestry Adjustment, Obesity

## Abstract

Polygenic risk scores (PRS) for type 2 diabetes (T2D) often lose accuracy when applied outside the populations in which they were developed. Partitioned PRS (PPRS) mitigate this by decomposing T2D risk into biologically interpretable pathways (e.g., obesity, body fat), but have not been adapted to Indonesians. We present PPRS-ID, an Indonesian-adjusted PPRS that integrates a locally derived obesity PRS with a population-specific linkage disequilibrium resource.

We analyzed 2,936 Indonesian participants to construct an Indonesian obesity PRS. To localize the T2D partitions, we harmonized SNPs with the published T2D PPRS and addressed limited direct overlap via LD proxy mapping. For this, we built a merged GRCh38 LD reference by liftover of West Javanese whole-genome sequences (n=227), performing per-chromosome QC and imputation against 1000 Genomes, and then merging the imputed West Java genomes with 1000 Genomes to form a combined panel. Signed LD correlations (r) were computed within 1 Mb of PPRS loci, enabling projection of T2D effects onto Indonesian obesity SNPs. Partition-specific hybrid weights were then formed by blending projected T2D betas with Indonesian obesity betas using biologically informed α parameters.

An ancestry analysis confirmed that Indonesian samples cluster distinctly from other 1000 Genomes groups, supporting the need for population-aware LD reference. Direct overlap between T2D PPRS and our obesity PRS comprised a single SNP; LD mapping recovered 10 additional proxies with r^2^ > 0.4. The evaluation on the UK Biobank Asian subset, PPRS-ID achieved an AUC of 0.633 for T2D discrimination. In a head-to-head test of the obesity pathway within Indonesians, the Indonesian obesity PRS outperformed the original obesity partition (AUC 0.594 vs. AUC 0.465).

PPRS-ID demonstrates a feasible path to population-tailored, pathway-aware T2D risk prediction in Indonesians. Ongoing work focuses on larger Southeast Asian validation, refined partition weighting, and assessment of clinical utility.

## 1. Background

Polygenic risk scores (PRS) have emerged as powerful tools for quantifying inherited susceptibility to complex diseases by aggregating the effects of numerous genetic variants across the genome [1,2]. In metabolic disorders such as type 2 diabetes (T2D), PRS enable stratification of individuals based on their genetic liability and hold promise for early risk prediction, personalized prevention, and population-level screening strategies. However, the clinical and predictive value of PRS remains heavily dependent on the ancestry of the population in which they are trained and applied. The majority of genome-wide association studies (GWAS) underlying current PRS are based on European-ancestry populations, which limits their transferability to genetically diverse groups such as Southeast Asians [3,4]. Discrepancies in allele frequencies, linkage disequilibrium (LD) patterns, and environmental exposures often result in reduced predictive accuracy and biased effect estimates when these scores are applied to other populations.

To improve interpretability and reduce overfitting, Suzuki et al. [5] recently introduced a multiancestry-weighted partitioned PRS (PPRS) for T2D, in which genome-wide genetic risk is decomposed into biologically meaningful pathways. This framework partitions variants according to their involvement in specific metabolic mechanisms—such as obesity and adiposity, insulin resistance, lipid metabolism, and glycaemic regulation—allowing disease risk to be contextualized through intermediate physiological processes. While this partitioned approach enhances interpretability and partially improves cross-population performance, it has not yet been adapted or validated in Southeast Asian populations. Considering the unique genetic diversity and admixture patterns across the Indonesian archipelago, which bridges East Asian and Oceanic ancestries, population-specific adaptation of the PPRS framework is both scientifically and clinically valuable.

To address this gap, we propose the Indonesian-adjusted Partitioned Polygenic Risk Score (PPRS-ID), a framework designed to tailor T2D risk prediction for the Indonesian population. PPRS-ID integrates a locally derived Indonesian obesity PRS, constructed from a cohort of 2,936 genotyped individuals [6], with the multi-pathway T2D PPRS developed by Suzuki et al. [5]. Recognizing that LD structure plays a critical role in PRS portability, we further introduce a population-specific LD reference panel by merging the 1000 Genomes Project Phase 3 reference [7] with West Javanese whole-genome sequencing (WGS) data [8]. This combined reference better captures regional haplotype variation and allele correlations, enabling LD proxy mapping and improved trans-ancestry SNP alignment.

By combining local effect sizes with globally informed pathway partitions, the PPRS-ID aims to improve both the accuracy and biological interpretability of T2D PRS in Indonesians. This work represents one of the first steps toward developing population-adjusted, pathway-aware PRS frameworks in Southeast Asia, contributing to the broader goal of equitable precision medicine. The following sections describe our methodology, data integration strategy, and preliminary findings on the predictive performance of PPRS-ID relative to existing models.

## 2. Methods

### 2.1 Dataset

The Indonesian dataset consisted of 2,936 individuals (959 males, 1,977 females) recruited between 2021 and 2024, with body mass index (BMI) as the primary phenotype. Genotyping was performed using the KalGen01 custom SNP array, designed to capture both common and rare variants relevant to Indonesian populations. Standard quality control steps included call rate filtering, Hardy–Weinberg equilibrium testing, and relatedness checks.

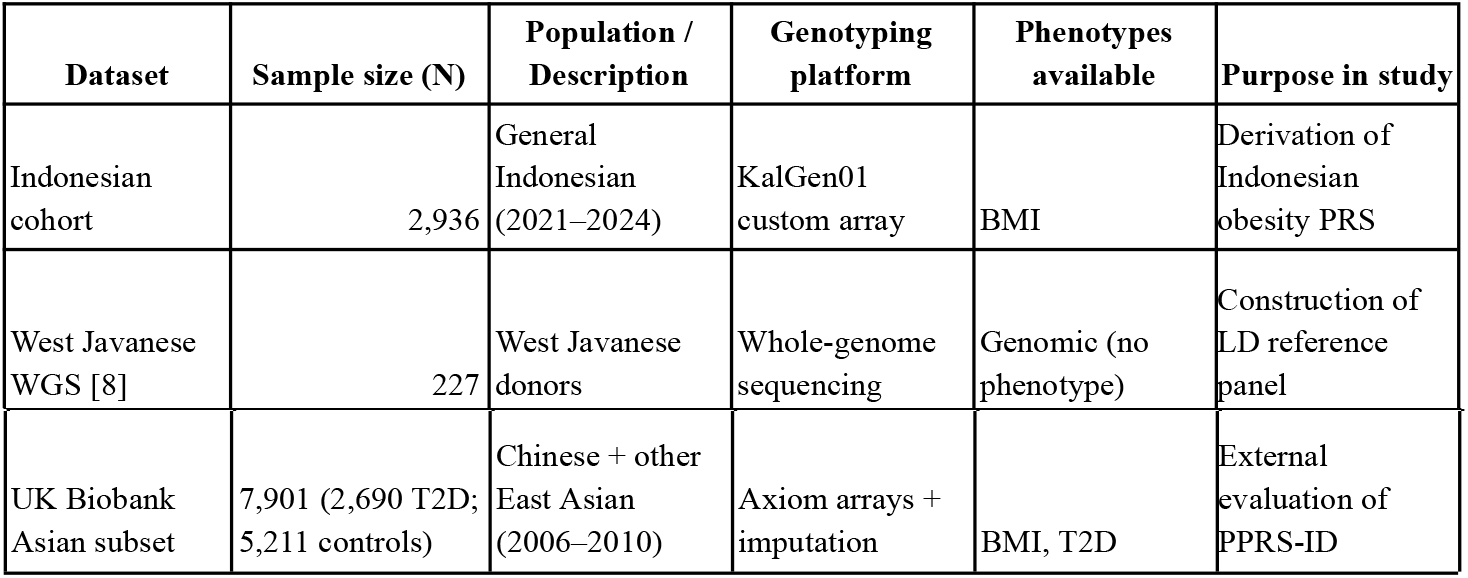

To build a population-specific LD reference, we incorporated 227 whole-genome sequences (WGS) from West Javanese donors published by Ardiansyah et al [8]. These sequences provided population-specific haplotype structure and novel variant representation, complementing the 1000 Genomes reference.

For external evaluation, we used the UK Biobank Asian subset (n = 7,901; 2,690 with T2D and 5,211 controls), consisting primarily of Chinese and other East Asian participants. BMI and T2D phenotype data were obtained from baseline assessments.

### 2.2 West Javanese Data Processing

The whole-genome sequences from West Javanese donors conducted by previous studies [8], were first harmonized to the same reference assembly used for downstream analyses. The raw WGS VCFs (originally on GRCh37) were liftovered to GRCh38, then compressed, sorted, and partitioned into autosomal chromosomes (1-22) to enable chromosome-wise processing. We applied targeted QC/cleanup to remove problematic sites and genotype calls (for example, records with missing alternate alleles or inconsistent genotype encodings) so that variant representation would be compatible with the 1000 Genomes reference and with the partitioned PRS framework mapped on GRCh38.

To recover missing genotypes and produce a population-representative LD resource, we performed genotype imputation on each chromosome using the 1000 Genomes Project as the reference panel and an appropriate GRCh38 genetic map. Imputation filled sparse or missing calls in the West Javanese WGS while preserving local haplotype structure. Imputed West Java chromosomes were then merged with the corresponding 1000 Genomes chromosomes to create a combined GRCh38 reference panel. Each merged chromosome file was indexed and prepared for downstream analyses. The resulting 1000G + West-Java panel provided a harmonized, population-aware LD reference that we used for LD proxy mapping and for computing signed LD correlations when projecting T2D effect sizes onto Indonesian obesity SNPs.

### 2.3 Partitioned PRS Construction and LD Mapping

An obesity PRS was first derived from the Indonesian cohort using genome-wide association analysis, using BMI as the continuous trait while also applying ancestry adjustment [5]. SNPs overlapping with Suzuki’s partitioned T2D PRS were harmonized on the GRCh38 build. Because overlap was limited, we expanded the SNP set via LD proxy mapping using a merged GRCh38 reference panel that included both the 1000 Genomes Project and WGS data from West Javanese individuals. Signed LD correlations (r) were computed within ±1 Mb windows, and effect sizes from T2D SNPs were projected onto obesity SNPs 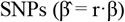.

Partition-specific hybrid weights were then generated by combining projected T2D betas with obesity-derived betas according to biologically informed α parameters (α=0.3 for obesity, α=0.7 for residual glycaemic pathways, α=0.3 for metabolic syndrome, α=0.3 for body fat). PRS scoring was performed in PLINK2, and the resulting PPRS-ID was evaluated in the UK Biobank Asian subset.

## 3. Results and Discussion

The ancestry analysis confirmed that Indonesian and West Javanese samples clustered distinctly compared to other 1000 Genomes populations, validating the need for a population-specific LD reference (Figure 2).

**Fig. 1.**
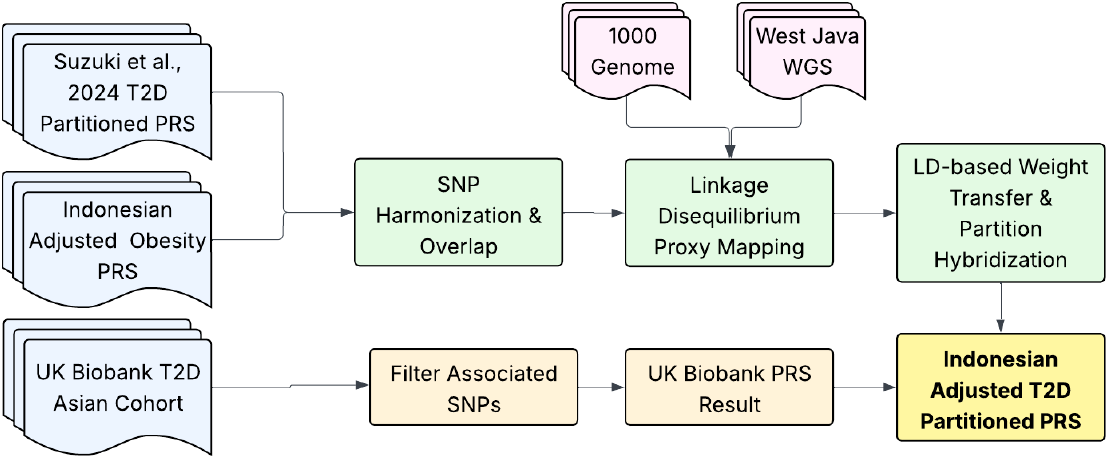
Workflow for constructing an Indonesian-adjusted partitioned PRS for T2D. Suzuki et al.’s partitioned PRS and a locally derived Obesity PRS were harmonized, then linked via LD mapping using a merged 1000 Genomes + West Java reference. Effect sizes were transferred and blended per partition to yield an Indonesian-adjusted partitioned PRS, which was subsequently validated against UK Biobank Asian cohorts.

**Fig. 2.**
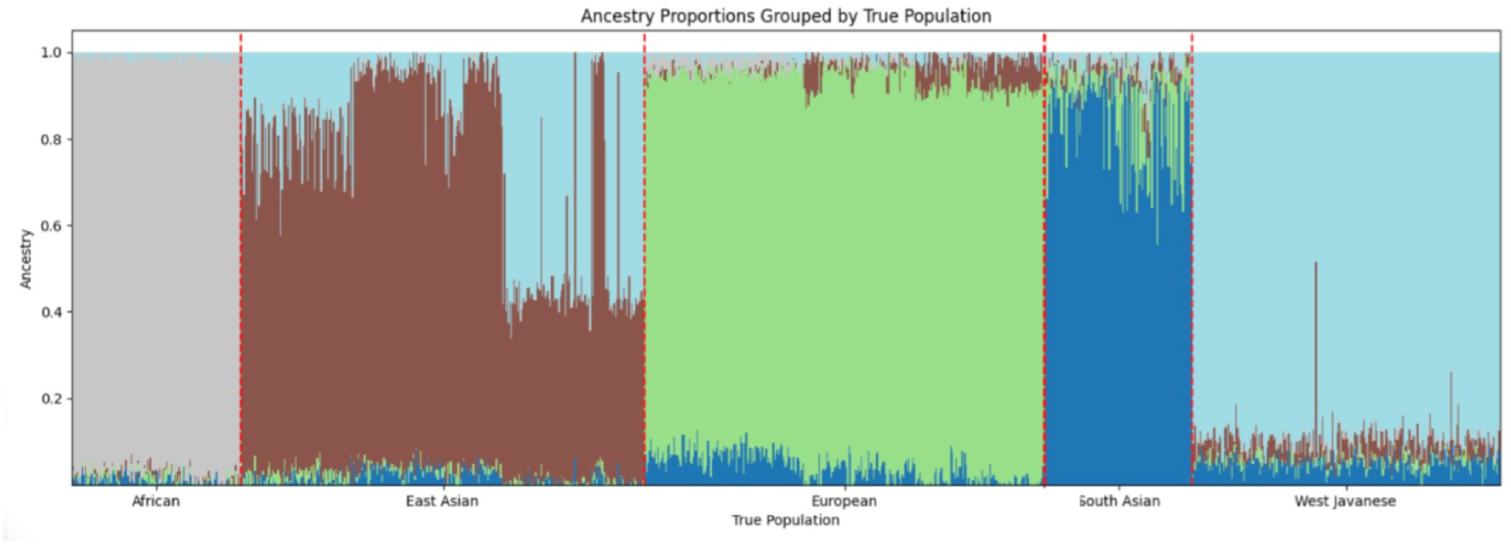
Ancestry proportions across global and West Javanese populations, showing distinct clustering of Indonesian (West Javanese) samples compared to 1000 Genomes groups.

We found 1 SNP overlap between Suzuki’s partitioned T2D SNPs and our Indonesian obesity PRS; however, the merged LD reference improved recovery of proxy SNPs across partitions by adding 10 matched SNPs with aligned R^2^ >0.4. Evaluation of PPRS-ID in the UK Biobank Asian subset yielded an AUC of 0.633 for discriminating T2D cases from controls (Figure 3). The distribution plots show a rightward shift in scores for cases compared to controls, demonstrating that transferability improves when pathway-specific weighting is combined with a locally trained component.

**Fig. 3.**
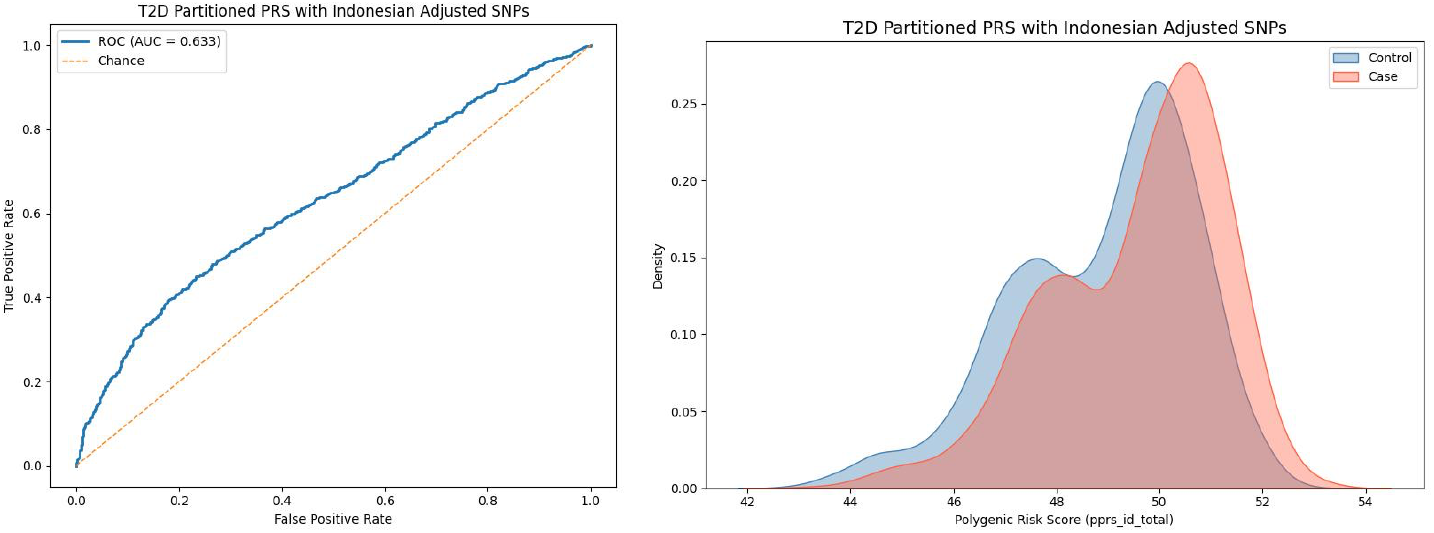
Performance of the Indonesian-adjusted partitioned PRS (PPRS-ID) for type 2 diabetes. (Left) ROC curve showing an AUC of 0.633 in the UK Biobank Asian subset. (Right) Distribution of PPRS-ID values, with T2D cases shifted toward higher scores compared to controls.

A head-to-head analysis of the obesity pathway PRS in Indonesians further highlighted the value of local effect sizes: the Indonesian-derived obesity PRS achieved AUC = 0.594, Sensitivity = 0.404, Specificity = 0.806, outperforming Suzuki’s original obesity PPRS (AUC = 0.465) when tested on the same cohort (Figure 4). The higher specificity of the Indonesian PRS suggests that it is more effective at correctly identifying non-obese individuals, which is important for minimizing false positives in clinical translation. While sensitivity remains modest, the overall improvement in discrimination emphasizes that local GWAS-derived betas capture relevant risk architecture not represented in external cohorts.

**Fig. 4.**
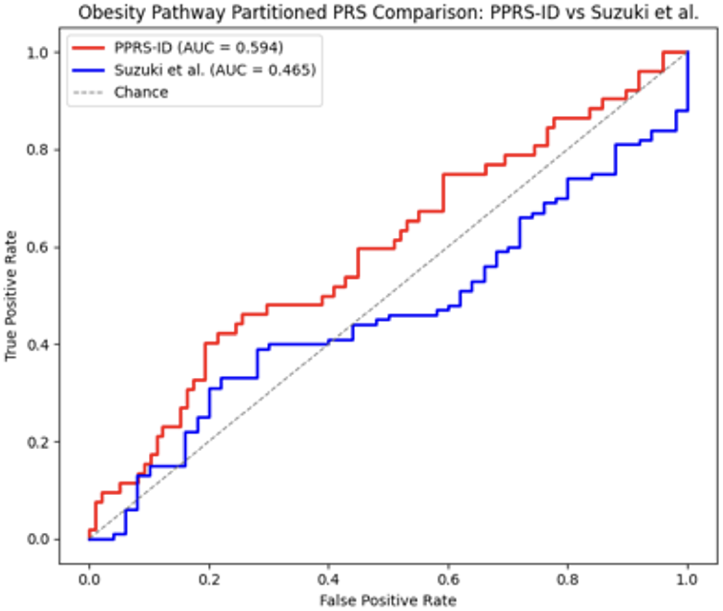
Head-to-head comparison of the obesity pathway PRS. The Indonesian-adjusted partitioned PRS (PPRS-ID, red) achieved better discrimination (AUC = 0.594) than Suzuki et al.’s obesity partition (blue, AUC = 0.465) when tested in the Indonesian cohort.

Overall, these findings demonstrate two key novelties: (i) embedding a locally trained obesity PRS into a global partitioned T2D framework, and (ii) constructing a population-aware LD reference by merging 1000 Genomes with West Javanese WGS data. Together, these innovations improve both portability and biological interpretability of PRS for T2D in underrepresented populations.

## 4. Conclusion

We present PPRS-ID, an Indonesian-adjusted partitioned PRS for T2D that integrates local obesity effect sizes with Suzuki’s PPRS architecture and leverages a merged 1000G+West Java LD reference. Preliminary results demonstrate moderate predictive accuracy (AUC 0.633 in UKBB Asians) and improved performance of the obesity partition in Indonesian data (AUC 0.594 vs 0.465). This study illustrates the feasibility of adapting partitioned PRS frameworks for underrepresented populations and underscores the importance of local genomic resources in precision medicine.

However, the current work is limited by the modest number of directly overlapping SNPs, reliance on proxy recovery via LD, and the relatively small size of available Indonesian and West Javanese datasets. These factors may constrain the stability and generalizability of effect size estimates. Future directions include validation in larger Southeast Asian cohorts, refinement of partition-specific weighting schemes, and exploration of clinical utility in personalized prevention and treatment strategies.

## Data Availability

All data produced in the present study are available upon reasonable request to the authors

